# Integrative transcriptomics analysis for uterine leiomyosarcoma identifies aberrant activation of cell cycle-dependent kinases and their potential therapeutic significance

**DOI:** 10.1101/2022.01.06.22268775

**Authors:** Kosuke Yoshida, Akira Yokoi, Tomofumi Yamamoto, Yusuke Hayashi, Jun Nakayama, Tsuyoshi Yokoi, Hiroshi Yoshida, Tomoyasu Kato, Hiroaki Kajiyama, Yusuke Yamamoto

**Affiliations:** Department of Obstetrics and Gynecology, Nagoya University Graduate School of Medicine, Nagoya, Japan; Institute for Advanced Research, Nagoya University, Nagoya, Japan; Laboratory of Integrative Oncology, National Cancer Center Research Institute, Tokyo, Japan; Department of Drug Safety Sciences, Division of Clinical Pharmacology, Nagoya University Graduate School of Medicine, Nagoya, Japan; Department of Diagnostic Pathology, National Cancer Center Hospital, Tokyo, Japan; Department of Gynecology, National Cancer Center Hospital, Tokyo, Japan

**Keywords:** uterine leiomyosarcoma, RNA-seq, cell cycle, PLK1, CHEK1,

## Abstract

**Purpose:** Uterine leiomyosarcoma is among the most aggressive gynecological malignancies. No effective treatment strategies have been established. This study aimed to identify novel therapeutic targets for uterine leiomyosarcoma based on transcriptome analysis and assess the preclinical efficacy of novel drug candidates.

**Experimental Design:** Transcriptome analysis was carried out using fresh-frozen samples of six uterine leiomyosarcomas and three myomas. The Ingenuity Pathway Analysis was then used to identify potential therapeutic target genes for uterine leiomyosarcoma. Moreover, our results were validated using three independent datasets, including 40 uterine leiomyosarcomas. Then, the inhibitory effects of several selective inhibitors for the candidate genes were examined using the SK-UT-1, SK-LMS-1, and SKN cell lines.

**Results:** We identified 512 considerably dysregulated genes in uterine leiomyosarcoma compared with myoma. The Ingenuity Pathway Analysis showed that the function of several genes, including CHEK1 and PLK1, were predicted to be activated in uterine leiomyosarcoma. Through an *in vitro* drug screening, PLK1 or CHEK1 inhibitors (BI 2536 or prexasertib) were found to exert a superior anti-cancer effect against cell lines at low nanomolar concentrations and induced cell cycle arrest. In SK-UT-1 tumor-bearing mice, BI 2536 monotherapy demonstrated a marked tumor regression. Moreover, the prexasertib and cisplatin combination therapy also reduced tumorigenicity and prolonged survival.

**Conclusion:** We identified the upregulated expression of *PLK1* and *CHEK1*; their kinase activity was considered to be activated in uterine leiomyosarcoma. BI 2536 and prexasertib demonstrate a significant anti-cancer effect; thus, cell cycle-related kinases may represent a promising therapeutic strategy for treating uterine leiomyosarcoma.

**Translational relevance:** The development of next-generation sequencing has had an immense impact on cancer research. However, the biological features of uterine leiomyosarcoma are not fully understood. Hence, no effective treatment strategies have been established based on its molecular background. In this research, we were able to assess the transcriptional profiles of 46 patients with uterine leiomyosarcoma using three independent datasets and through the assistance of our cohort. The integrative transcriptional analysis showed that the upregulation and activation of cell cycle-related genes were the dominant features of uterine leiomyosarcoma. Afterward, we demonstrated that PLK1 or CHEK1 inhibition induced cell cycle arrest and caused DNA damage, which resulted in cell death in leiomyosarcoma-derived cell lines. Moreover, these drugs had a more significant anti-cancer effect in the mice model. These data suggest that cell-cycle-dependent kinases represent novel therapeutic targets and could potentially improve the outcome for patients with uterine leiomyosarcoma.

## Introduction

Uterine sarcomas are a rare subset of gynecologic malignancies with an extremely aggressive behavior. Uterine sarcomas are further classified into one of three groups: leiomyosarcoma (LMS), endometrial stromal sarcoma, and adenosarcoma, of which LMS is the most common subtype (1,2). The annual incidence of uterine LMS (ULMS) is approximately 0.86 per 100,000 women, and the majority of the patients are postmenopausal (3,4). Complete surgical resection followed by adjuvant chemotherapy or radiation would be one of the reasonable management strategies for patients with early-stage disease, although most patients eventually experience a recurrence (1,2). The combination of docetaxel and gemcitabine has been widely used for patients with metastasis and seems to be partially effective (2). However, over the last few decades, the median overall survival (OS) of patients with metastatic ULMS has only been one or two years (4,5). Recently, novel agents, such as trabectedin, pazopanib, and eribulin, have been approved for soft-tissue sarcomas. Despite high expectations for these agents, the prognosis of patients with ULMS has not considerably improved (6-8). For example, a subgroup analysis showed that the median progression-free survival (PFS) and OS for ULMS patients treated with trabectedin were 4.0 and 13.4 months, respectively (6). Similarly, other clinical trials that evaluated pazopanib and eribulin for the treatment of ULMS showed a median OS of 17.5 and 12.7 months, respectively (7,8). Therefore, the clinical outcome of ULMS remains unsatisfactory, and new therapeutic agents are urgently needed.

Recently, the development of next-generation sequencing has enabled the genomic landscape to shed light on a variety of cancers. Several reports have revealed that alterations affecting *TP53, RB1, ATRX*, and *PTEN* frequently occur in ULMS (9-12). Moreover, in some cases, fusion genes, such as TNS1-ALK, ACTG2-ALK, and KAT6B-KANSL1, have been identified (12,13). Therefore, the genomic features of ULMS may be responsible for its aggressiveness. Additionally, gene expression profiles provide important information for comprehending cancer biology. However, only a few small-scale studies have been carried out at the RNA level in ULMS because RNA is less stable than DNA, and samples are rare (12,14). In one study, Aurora A and B kinases were identified as potential therapeutic targets in ULMS; however, a subsequent phase II study failed to demonstrate the single-agent activity of the Aurora kinase inhibitor, alisertib (14,15). Therefore, the development of new therapeutic agents for ULMS remains a challenge.

In the present study, we identified that cell cycle-related genes were upregulated and that their kinase activity was predictively activated in ULMS compared with myoma and normal myometrium. With three datasets and our cohort, this is one of the largest research projects assessing the transcriptional landscape of ULMS. Moreover, subsequent analyses showed that PLK1 and CHEK1 inhibition strongly induced cell cycle arrest and exerted superior anti-cancer effects both *in vitro* and *in vivo*.

## Materials and methods

### Patients

Archival fresh-frozen tumor samples stored at the National Cancer Center Biobank (Tokyo, Japan) were used. Since 2011, there have been six patients with ULMS who underwent surgery without neoadjuvant therapy. The sarcoma and adjacent myometrium tissues of the six patients were obtained. Moreover, three patients with benign leiomyoma were included as controls. The study protocol was approved by the ethics committee at our institution (approval No. 2020-160). We obtained written informed consent from all patients.

### RNA extraction and transcriptome analysis

Total RNA was extracted from six ULMS and three myoma samples using the miRNeasy Mini Kit (Qiagen, Hilden, Germany), and pair-end sequencing was carried out using a DNBSEQ-G400 (MGI Tech, Shenzhen, China) by Azenta (South Plainfield, NJ). From the sequencing data, expression levels for each gene were quantified by Kallisto (16). Then, the data were summarized using the tximport package (ver. 1.18.0) of R software (ver. 4.0.3) and Rstudio (RStudio, Boston, MA), and scaledTPM counts were used for further analysis. Excluding genes with low read coverage (maximum read count: < 100 reads), 3,070 differentially expressed genes (DEGs, |log2FC| > 1) between the ULMS and myoma samples were used for a heatmap analysis. The heatmap.2 function of the gplots package (ver. 3.1.0) was used after the data were converted to base 10 logarithms and z-scores. For volcano plots, the adjusted *P* values for each gene were calculated by the Wald test in DESeq2 (ver. 1.30.0) using data for 23,353 annotated genes. Subsequently, we performed pathway and upstream regulator analysis by Ingenuity Pathway Analysis (IPA, Qiagen) using the significant DEGs identified on the volcano plot.

### NCBI GEO dataset

The three datasets, GSE36610 (12 ULMS and ten myometrium samples), GSE64763 (25 ULMS, 25 myomas, and 29 myometrium samples), and GSE68295 (three ULMS, three myomas, and three myometrium samples), were downloaded from the NCBI GEO database. The three datasets were microarray-based transcriptional profiles, and 10,641 common gene symbols were used for analysis (Supplementary Fig. S1A). The expression data were converted to z-scores, and 1,683 DEGs (the difference between the mean z-scores of ULMS and myometrium is greater than 1) were used to generate the heatmap and principal component analysis (PCA). The PCA was visualized using the prcomp and plot3d functions of the rgl package (ver. 0.100.54). For volcano plot, log2FC and adjusted *P* values for each gene were calculated for each dataset. Then, pathway analysis was performed by IPA using the common DEGs identified on the volcano plots.

### Cell lines

Three ULMS-derived cell lines, SKN, SK-UT-1, and SK-LMS-1, were used. SKN was purchased from the Japanese Cancer Research Resources Bank (Osaka, Japan), and SK-UT-1 and SK-LMS-1 were purchased from the American Type Culture Collection (Manassas, VA). SKN cells were maintained in Ham’s F12 medium (Sigma-Aldrich, St. Louis, MO) containing 10% fetal bovine serum (Thermo Fisher Scientific, Waltham, MA) and antibiotics. SK-UT-1 and SK-LMS-1 cells were maintained in MEM (Nacalai Tesque, Kyoto, Japan) containing 10% fetal bovine serum, 1 mM sodium pyruvate (Thermo Fisher Scientific), and antibiotics. The cell lines tested negative for mycoplasma contamination and were used between 5 and 40 passages for experiments.

### Chemicals

All selective kinase inhibitors were purchased from Selleck (Houston, TX), and their putative targets are shown in Supplementally Table S1. Briefly, BI 2536 and volasertib (BI 6727) are PLK1 inhibitors. Prexasertib HCl (LY2606368) and PF-477736 are ATP-competitive CHEK1/2 inhibitors. Dinaciclib (SCH727965) and flavopiridol (Alvocidib) are pan-CDK inhibitors. AT9283 is a JAK2/3 and Aurora A/B inhibitor, and tozasertib (MK-0457) is a pan-Aurora inhibitor. JNJ-7706621 is a pan-CDK and potent Aurora A/B inhibitor, and BAY 1217389 is a TTK inhibitor. Pazopanib is an approved multi-target inhibitor. The drugs were dissolved in DMSO as stock solutions and further diluted in the culture medium for experiments. Moreover, cisplatin was purchased from Nichi-Iko Pharmaceutical (Toyama, Japan).

### Small interfering RNAs (siRNAs)

Silencer Select Pre-designed siRNAs for each gene (Thermo Fisher Scientific) were used, and the assay IDs were as follows; s448 (siPLK1 No. 1), s449 (siPLK1 No. 2), s503 (siCHEK1 No. 1), s504 (siCHEK1 No. 2), s22119 (siCHEK2 No. 1), and s22121 (siCHEK2 No. 2). Cells were transfected with 3 nM siRNA using Lipofectamine RNAi Max (Thermo Fisher Scientific).

### Cell viability assay

Cells were seeded into 96-well plates. Immediately after attachment, the cells were treated with the inhibitors and incubated for 72 h. For siRNAs, cells were transfected with the siRNAs and incubated for 24, 48, and 72 h. Cell viability was assessed using the CellTiter-Glo 2.0 Cell Viability Assay (Promega, Madison, WI), and the luminescence measurements were taken 10 min after adding the reagent using a microplate reader (Molecular Devices, San Jose, CA). Viability was calculated with the percentage of untreated cells, and experiments were performed in triplicate and repeated three times. IC50 and drug dose-response curves were calculated in GraphPad Prism 7 (Version 7.0d, GraphPad Software, San Diego, CA). Synergy was determined using CompuSyn software (version 1.0) (http://www.combosyn.com/index.html).

### Cell cycle assay

Cells were seeded in 6-well plates and grown to approximately 80% confluency. Then, cells were treated with selective inhibitors for 24 h. The cells were harvested following trypsinization, washed with 3% FBS/PBS, and fixed in cold 70% ethanol. The cells were then resuspended in 3% FBS/PBS and stained with ReadiDrop Propidium Iodide (Bio-Rad Laboratories, Hercules, CA). Cell analyzer EC800 (Sony, Tokyo, Japan) was used for the analysis. The resulting data were analyzed with FlowJo software (BD Biosciences). Experiments were performed in triplicate and repeated three times.

### qRT-PCR

Paired ULMS and adjacent normal tissues were used, and total RNA was extracted as described previously. Total RNA was extracted from transfected cells, and cDNA was synthesized using SuperScript III Reverse Transcriptase (Thermo Fisher Scientific). The QuantiTect SYBR Green PCR Kit (Qiagen) or THUNDERBIRD SYBR qPCR Mix (Toyobo, Osaka, Japan) were used. Specific primers were synthesized by Fasmac (Kanagawa, Japan), and the primer sequences are shown in Supplementary Table S2. The amplification program was as follows: denaturation at 95°C for 10 min, followed by 40 amplification cycles of 95°C for 15 s and 60°C for 60 s. The amplified product was monitored by measuring SYBR Green I dye fluorescence intensity, and β-actin was used as a reference gene to normalize expression.

### Western blot analysis

Cells were treated with prexasertib for 16 h, and then, total protein extracts were prepared using the M-PER Mammalian Protein Extraction Reagent (Thermo Fisher Scientific) containing the Halt Protease & Phosphatase Inhibitor Single-Use Cocktail (Thermo Fisher Scientific). After quantification, 10 μg of total protein was separated on Mini-PROTEAN TGX gels (4%–20%, Bio-Rad Laboratories) and transferred to PVDF membranes. The following primary antibodies were used: Chk1 (2G1D5) Mouse mAb #2360 (Cell Signaling Technology), Phospho-Chk1 (Ser296) Antibody #2349 (Cell Signaling Technology), and β-actin (C4) (MAB1501, Merck, Germany). Horseradish peroxidase-conjugated anti-mouse IgG (NA931) and horseradish peroxidase-conjugated anti-rabbit IgG (NA934) were purchased from GE Healthcare (Buckinghamshire, UK). Protein bands were visualized using ImmunoStar LD (Fujifilm Wako Pure Chemical, Osaka, Japan) and ImageQuant LAS-4000 (Fujifilm, Tokyo, Japan).

### Immunofluorescence

After incubation with a prexasertib-containing medium for 24 h, cells were fixed with 4% paraformaldehyde. The cells were then treated with 0.3% Triton-X/Blocking One solution (Nacalai Tesque). Phospho-Histone H2A.X (Ser139) Antibody #2577 (Cell Signaling Technology, Danvers, MA), Alexa-Fluor 488 Goat anti-Rabbit IgG (H+L) Cross-Absorbed Secondary Antibody (Thermo Fisher Scientific), and Hoechst 33342, trihydrochloride, trihydrate (Thermo Fisher Scientific) were used for detection and staining, and images were captured using a BZ-X700 fluorescence microscope (Keyence, Osaka, Japan).

### Animal studies

All mouse experiments were approved by the National Cancer Center Research Institute, Institute of Laboratory Animal Research (Number: T18-009). Four-week-old female BALB/C nude mice were used for the animal experiments, and 3.0 × 10^6^ SK-UT-1 cells were injected into the right flank of the mice. BI 2536 was dissolved in hydrochloric acid (0.1N) and diluted with 0.9% NaCl. Prexasertib was dissolved in a vehicle (5% DMSO + 40% PEG 300 + 5% Tween80 + ddH2O), and cisplatin was diluted with 0.9% NaCl. The drugs were administered intraperitoneally twice a week. Mice were monitored carefully, and tumor volume was calculated using the modified ellipsoid formula (Length × Width^2^ × 0.5). Mice were sacrificed when the tumors reached a volume of 2,000 mm^3^.

### Statistical analysis

Statistical analysis was performed with RStudio and R software (ver. 4.0.3). Welch’s *t*-test was used to determine the significance of differences between the means of two sets of data. Paired t-test was used to determine the significance of differences between the paired LMS and myometrium samples. Dunnett’s test was used for multiple comparisons with a control group using the multicomp package (ver. 1.4-17). Kaplan-Meier curves and a log-rank test were used for the analysis of the survival. A *P* value of less than 0.05 was considered statistically significant.

### Data availability statement

The data generated in this study are publicly available in Gene Expression Omnibus (GEO) at GSE185543.

## Results

### Transcriptome analysis of clinical samples

Transcriptome analysis was performed using six ULMS and three myoma samples. The median age of the patients with ULMS was 59.5 (range, 53–79) years and all patients underwent surgery without neoadjuvant therapy. The heatmap showed that the gene expression profile of ULMS was quite different from that of myoma (Fig. 1A). The expression of 23,353 genes was compared by multivariate analysis. There were 387 significantly upregulated and 125 significantly downregulated genes in ULMS based on a cut-off of |log2FC| > 1 and an adjusted *P*-value < 0.05 (Fig. 1B and Supplementary Table S3 & S4). To assess the putative function of the 512 DEGs, pathway analysis was performed using the IPA software, which revealed that several pathways associated with the cell cycle and DNA damage checkpoint were significantly dysregulated. For example, these included “Kinetochore Metaphase Signaling Pathway (*P* = 5.01E-24),” “Mitotic Roles of Polo-Like Kinase (*P* = 1.58E-11),” and “Cell Cycle: G2/M DNA Damage Checkpoint Regulation (*P* = 2.51E-7)” (Fig.1C). In addition, upstream regulator analysis using IPA revealed that the function of CDK1, AURKB, PLK1, CHEK2, CHEK1, CDK2, and PRKDC was significantly activated in ULMS (Fig.1C and Table 1).

**Fig. 1.**
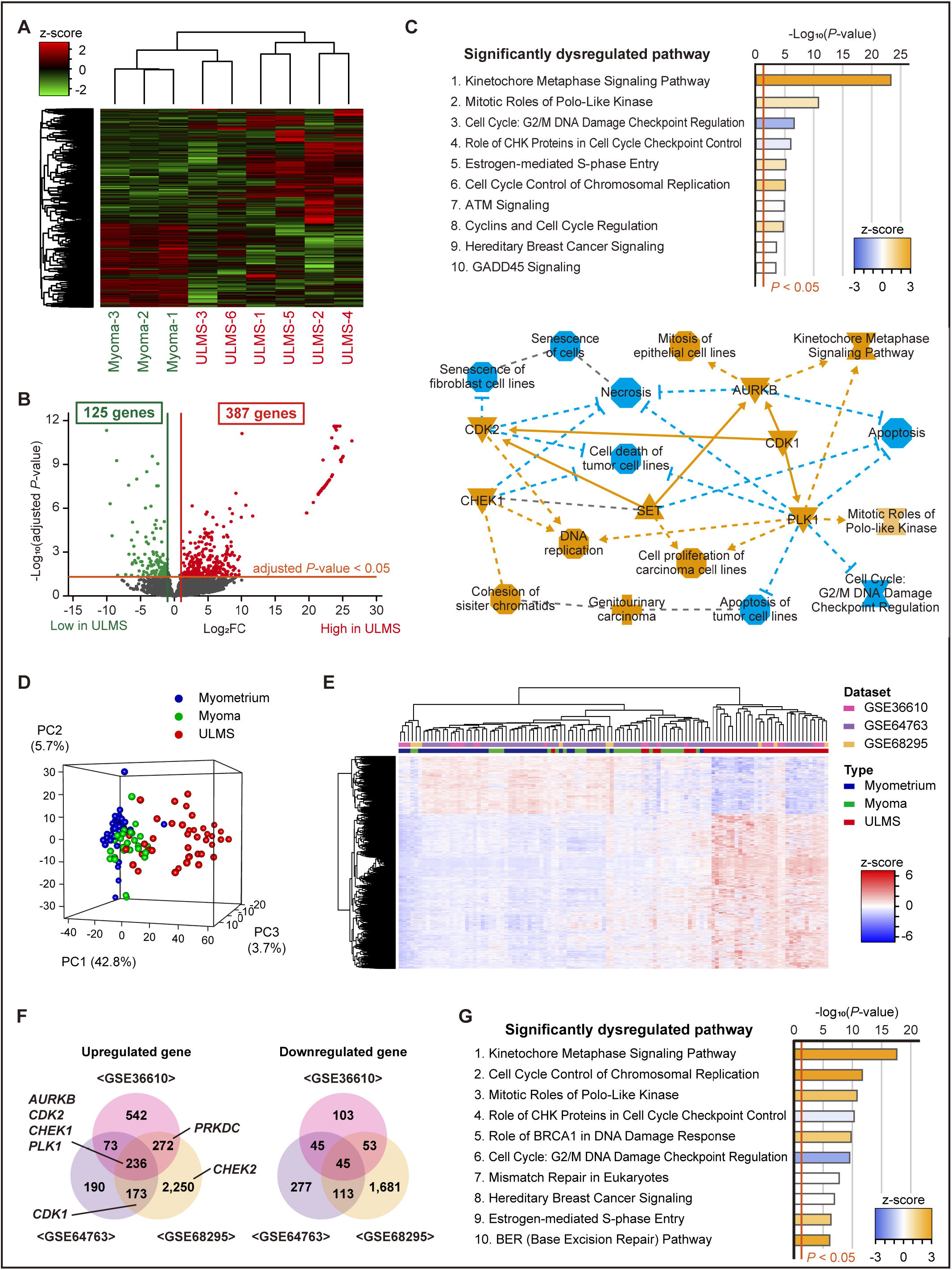
The transcriptome analysis of uterine leiomyosarcomas (LMSs) and myomas. **(A)** The hierarchical clustering and heatmap showing 3,070 differentially expressed genes (DEGs) between the ULMSs and myomas. The DEGs were defined as an absolute log2 fold change exceeding 1. **(B)** The volcano plot showing significant DEGs between LMSs and myomas. The adjusted *P* values for each gene were calculated by the Wald test in DESeq2. **(C)** The top ten significantly dysregulated pathways and the graphical summary based on Ingenuity Pathway Analysis (IPA) for the significant DEGs. The orange and blue nodes represent the activated and inhibited genes or pathways, respectively. The orange arrows and blue inhibitory arrows indicate activation and suppression, respectively. **(D)** The principal component analysis **(E)** The hierarchical clustering and heatmap for GSE36610, GSE64763, and GSE68295 dataset. Each data was converted to z-scores and merged. Comparing the mean expression of ULMS and myometrium, 1,683 DEGs were used for analyses. **(F)** The Venn diagram showing the significantly upregulated and downregulated genes between ULMS and myometrium in each dataset. The name of seven predictively activated upstream regulators in ULMS is shown. **(G)** The top ten significantly dysregulated pathways for the three datasets. The IPA analysis was performed using the 282 significant DEGs in common.

**Table 1.** The list of upstream regulators and their target molecules.

| Upstream Regulator (kinase) | Predicted Activation State | Target Molecules in Dataset | *P* value of overlap |
| --- | --- | --- | --- |
| CDK1 | Activated | AURKB, BIRC5, BUB1, BUB1B, CDC20, CDC25A, CDC25C, CDC6, CDT1, FEN1, FOXM1, GAS2L3, H2AX, MCM4, PBK, PLK1, PTTG1, SGO1, STMN1, TTK | 7.63E-10 |
| AURKB | Activated | AURKB, BIRC5, CDCA8, CENPA, DTL, KIF2C, SGO2, SKA1, SKA3, TOP2A | 6.24E-09 |
| PLK1 | Activated | BIRC5, BUB1, BUB1B, CCNB1, CDC20, CDC25C, CEP55, FBXO5, GTSE1, H2AX, HAUS8, KIF2C, KIFC1, PTEN | 2.48E-08 |
| CHEK2 | Activated | BIRC5, CDC25A, CDC25C, CDK1, FOXM1, TTK | 4.84E-05 |
| CHEK1 | Activated | CDC25A, CDC25C, CHEK1, CLSPN, E2F7, E2F8, H2AX | 1.46E-04 |
| TTK |  | BUB1, BUB1B, KNL1, RMI2 | 1.53E-04 |
| CDK2 | Activated | CCNA2, CDC25A, CDC6, CDK1, CDT1, CHEK1, KIF11, MCM4, MYBL2, PTTG1 | 5.66E-04 |
| CDKN1A |  | CDC6, CDK1, CDT1, CHEK1, H2AX | 8.10E-04 |
| WEE1 |  | CDK1, H2AX, STMN1 | 0.002 |
| PRKDC | Activated | CBX5, CHEK1, H2AX, PLK1, TPX2 | 0.002 |
| PLK3 |  | CDC25C, H2AX, PTEN | 0.003 |
| MAPK9 |  | CBX7, CCND3, CDC25A, CDC25C, CHEK1, H2AX | 0.004 |
| CCNB1 |  | BIRC5, BUB1, CDC25A, PBK | 0.006 |
| RPS6KA3 |  | CHEK1, H2AX, NEK2, SHANK3, STMN1 | 0.009 |
| BUB1B |  | BUB1B, CDC20 | 0.013 |
| RPS6KA2 |  | CHEK1, FBXO5, NEK2 | 0.016 |

Then, to validate our results, we used three GEO datasets, which were made up of the expression of 10,641 genes in 40 ULMS, 28 myomas, and 42 normal myometrium samples (Supplementary Fig. S1A). The PCA and heatmap analysis showed that the transcriptional profile of ULMS is different from that of myoma and normal myometrium (Fig.1D&1E). Then, comparing ULMS and myometrium samples in each dataset, we identified that 236 and 45 genes were commonly upregulated and downregulated in ULMS, respectively (Fig. 1F and Supplementary Fig. S1A). Moreover, the expression of the seven upstream regulators, which were identified in our cohorts, were confirmed to be upregulated in ULMS (Fig. 1F). Then, the IPA analysis using 281 DEGs validated the activation of “Kinetochore Metaphase Signaling Pathway (*P* = 2.58E-18)” and inhibition of “Cell Cycle: G2/M DNA Damage Checkpoint Regulation (*P* = 2.82E-10)” (Fig. 1G). Therefore, aberrant cell cycle regulation would be a dominant transcriptional feature of ULMS.

### In vitro screening of selective inhibitors for the activated upstream regulators

We considered the upregulated and activated key regulators as potential therapeutic targets for ULMS. Hence, the anti-cancer effects of selective inhibitors for these genes were evaluated using three cell lines derived from ULMS. First, we evaluated the efficacy of pazopanib, which is an approved drug for malignant soft-tissue tumors. Cells were treated with pazopanib for 72 h, and the IC50 values for SK-UT-1, SK-LMS-1, and SKN cells were 30.7, 62.8, and 5.7 μM, respectively (Supplementary Fig. S2A). Then, we assessed the efficacy of selective inhibitors for the target genes, and most of them were highly effective compared with pazopanib (Fig. 2A&2B and Supplementary Fig. S2B). The inhibitors for PLK1 (BI 2536 and volasertib) exhibited cytotoxicity at a lower nanomolar concentration in SK-UT-1 and SK-LMS-1 cells (Fig. 2A). Moreover, CHEK1/2 inhibitor (prexasertib and PF-477736) also showed a higher sensitivity in SK-UT-1 cells, with an IC50 below 10 nM (Fig. 2B). On the other hand, both BI 2536 and prexasertib were less effective in SKN cells compared with SK-UT-1 cells, although their effect was at least ten times greater than that of pazopanib.

**Fig. 2.**
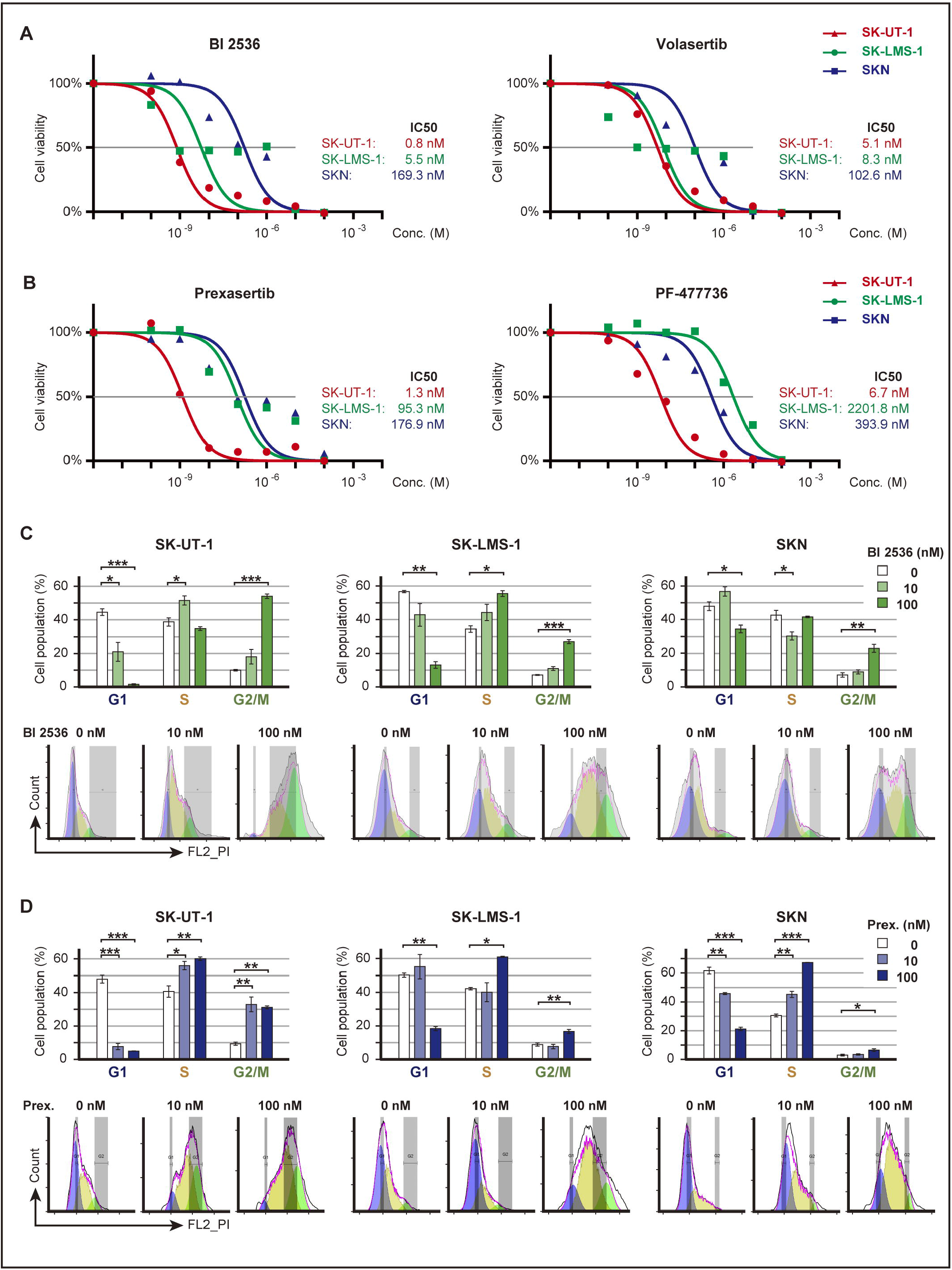
The inhibitory effects of PLK1 or CHEK1/2 inhibitors. **(A)** The effect of PLK1 inhibitors; BI 2536 and volasertib. **(B)** The effect of CHEK1/2 inhibitors; prexasertib and PF-477736. Cells were treated with each inhibitor for 72 h. Red, green, and blue represent SK-UT-1, SK-LMS-1, and SKN, respectively. Experiments were performed in triplicate and repeated three times, and IC50 and drug dose-response curves were calculated in GraphPad Prism 7. **(C)** Cell-cycle distribution of BI 2536-treated cells. **(D)** Cell-cycle distribution of prexasertib-treated cells. Cells were treated with each concentration of inhibitor for 24 h. Cell-cycle distribution was calculated by FlowJo, and the percentage of cells was compared using Dunnett’s test. Error bars represent standard errors of the mean.

Then, we evaluated the effect of BI 2536 and prexasertib on the cell cycle. In the 24 hour-treatment, 10 nM BI 2536 showed little effect, but 100 nM BI 2536 considerably decreased the cell population in the G1 phase and increased that in the S and G2/M phase (Fig. 2C). Similarly, when treated with 100 nM prexasertib, the cell population in the G1 phase was considerably decreased, whereas that in the S and G2/M phases was considerably increased in all cell lines (Fig. 2D). In particular, SK-UT-1 cells were highly sensitive to prexasertib, and 10 nM prexasertib was enough to induce cell cycle arrest.

### The effect of PLK1 inhibition

According to the results of integrative transcriptome analysis and drug screening, PLK1 is the most attractive therapeutic target. In our cohort, the expression of *PLK1* was significantly increased in ULMS compared with adjacent normal myometrium (*P* < 0.001, Fig. 3A). Thus, to assess the role of PLK1 in ULMS, we performed gene silencing experiments using siRNAs. Two siRNAs for PLK1 reduced the expression of *PLK1* to about 30–40% and increased the number of the round shape cells (Fig. 3B&3C). The cell cycle analysis showed that siRNAs for PLK1 considerably decreased the cell population in the G1 phase and increased that in the S and G2/M phase in all cell lines (Fig. 3D). Therefore, PLK1 knockdown significantly decreased the cell proliferation (in SK-UT-1, SK-LMS-1, and SKN; *P* < 0.01, *P* < 0.001, and *P* < 0.01, respectively, Fig. 3E). In particular, the effect of 3 nM siRNA was almost the same as that of 100 nM BI 2536 and caused complete growth arrest in SK-LMS-1 cells (Fig. 3D&3E).

**Fig. 3.**
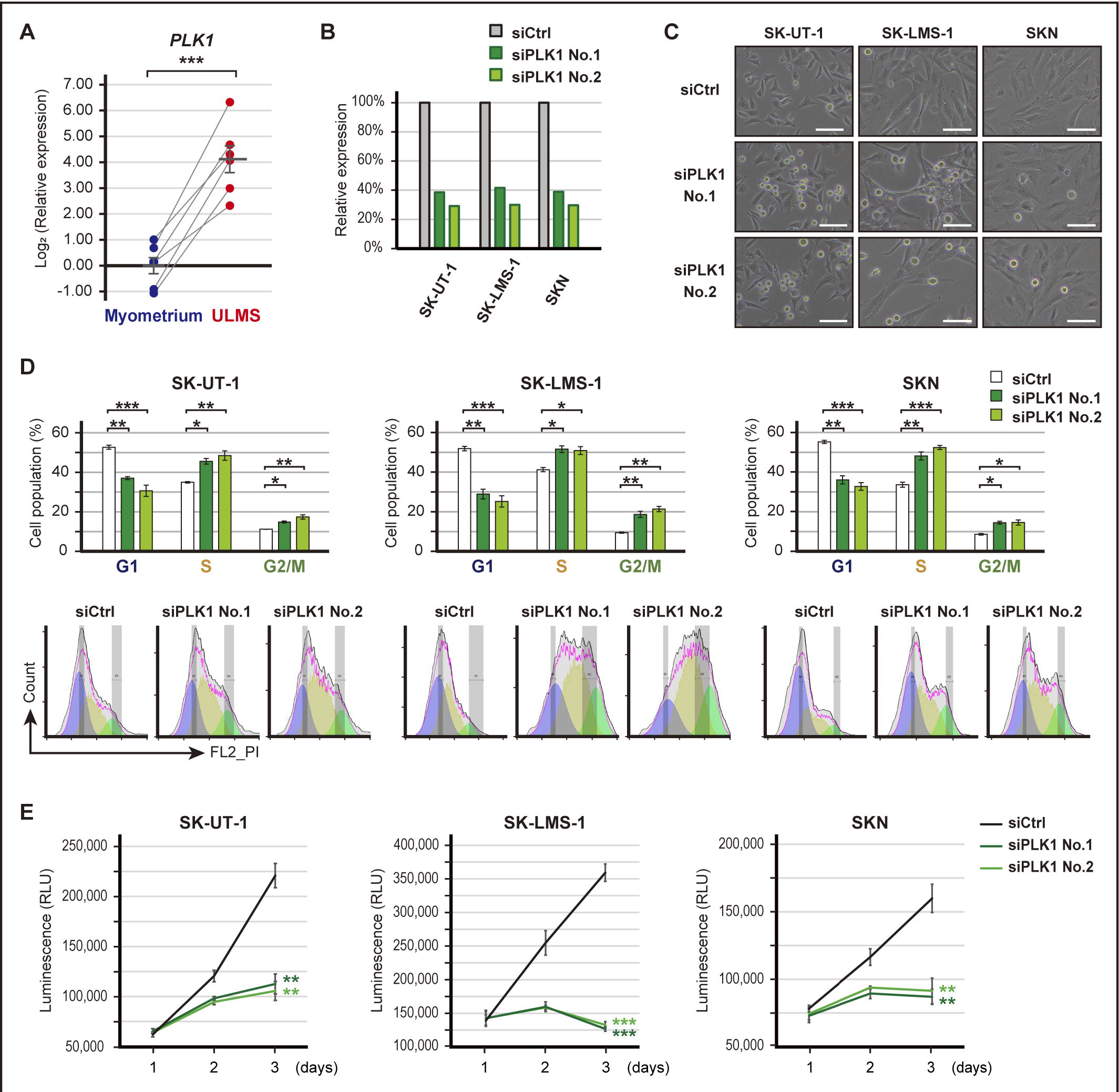
The effects of PLK1 silencing. **(A)** The relative expression of *PLK1* in paired ULMS and myometrium. The relative expression was compared using paired *t*-test. **(B)** Validation of *PLK1* suppression following transfection with 3 nM of siRNA for PLK1 (siPLK1). **(C)** The representative images of siPLK1 transfected cells. Scale bars show 100 μm. **(D)** Cell-cycle distribution of siPLK1 transfected cells. Cell-cycle distribution was calculated by FlowJo, and the percentage of cells was compared using Dunnett’s test. **(E)** The proliferation of siPLK1 transfected cells. Cell viability was measured at 24, 48, and 72 h, and the luminescence was compared using Dunnett’s test. Error bars represent standard errors of the mean, \**P* < 0.05, \*\**P* < 0.01, and \*\*\**P* < 0.001.

### The effect of CHEK1 inhibition

In addition to PLK1 inhibition, CHEK1/2 inhibition is also a promising therapeutic strategy to impair DNA damage response. Both *CHEK1* and *CHEK2* were upregulated in ULMS compared with adjacent normal myometrium (*P* < 0.01 and *P* < 0.01). The fold change of CHEK1 is larger than that of CHEK2 (Fig. 4A). The siRNA-mediated downregulation of *CHEK1* and *CHEK2* was confirmed by qRT-PCR, but the effect of siRNAs was different depending on the cell types (Fig. 4B). In SK-UT-1 cells, siRNAs for CHEK1 significantly inhibited cell proliferation (*P* < 0.001), whereas siRNAs for CHEK2 showed no effect on proliferation (Fig. 4C). In SK-LMS-1 and SKN cells, siRNAs for both CHEK1 and CHEK2 slightly but significantly inhibited the proliferation (Fig. 4C). Moreover, prexasertib decreased the expression of pCHEK1(Ser296) in a dose-dependent manner (Fig. 4D). Therefore, CHEK1 was more responsible for the prexasertib-induced growth arrest.

**Fig. 4.**
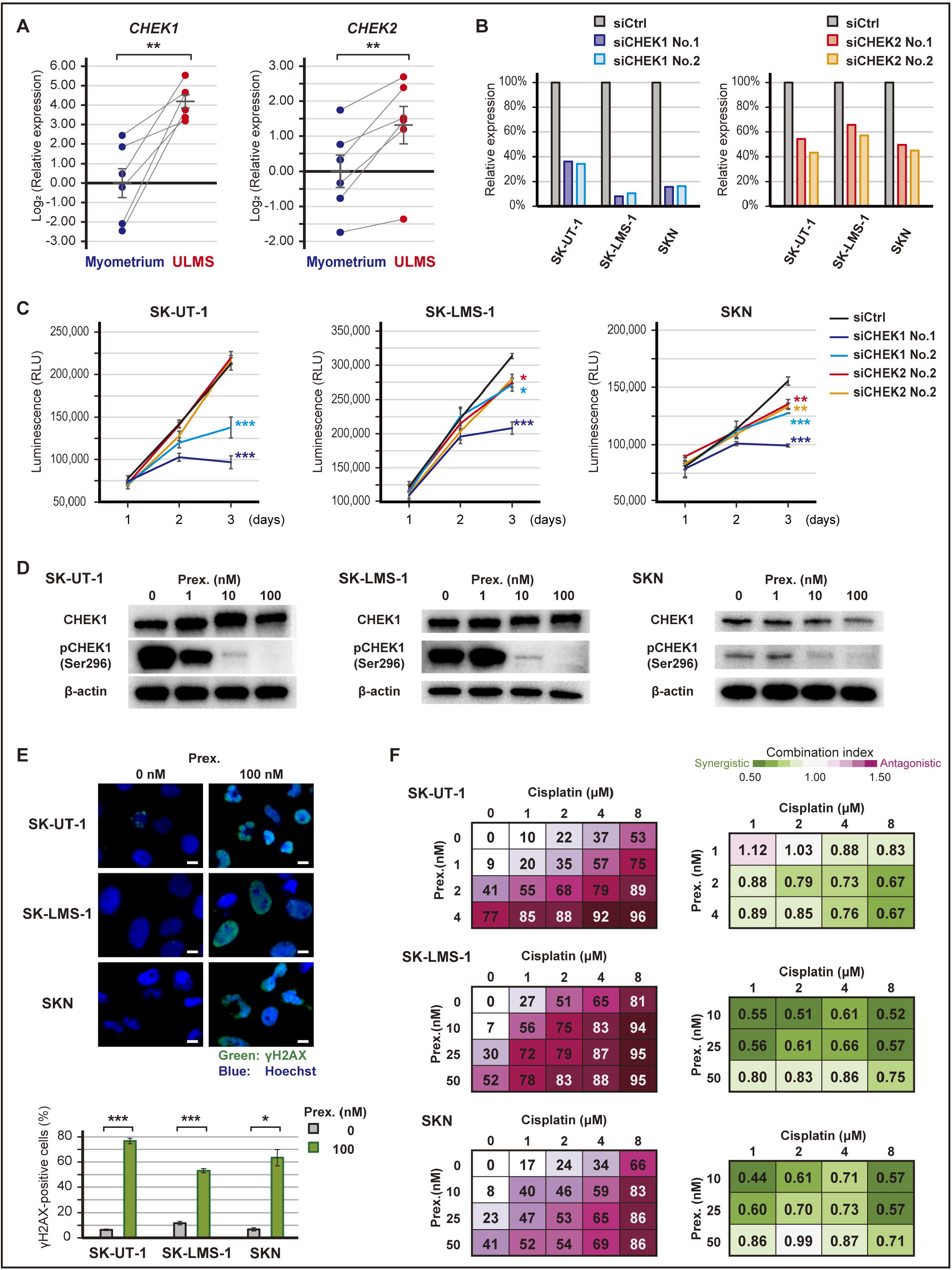
The effects of CHEK1 silencing and prexasertib. **(A)** The relative expression of *CHEK1* and *CHEK2* in paired ULMS and myometrium. The relative expression was compared using paired *t*-test. **(B)** Validation of *CHEK1* or *CHEK2* suppression following transfection with 3 nM of siRNA for CHEK1 (siCHEK1) or CHEK2 (siCHEK2). **(C)** The proliferation of siCHEK1 transfected cells. Cell viability was measured at 24, 48, and 72 h, and luminescence was compared using Dunnett’s test. **(D)** The expression of CHEK1 and pCHEK1(Ser296) protein in prexasertib-treated cells. Cells were treated with each concentration of prexasertib for 16 h. **(E)** Immunofluorescent of γH2AX in prexasertib-treated cells. Cells were treated with 0 or 100 nM prexasertib for 24 h, and the percentage of γH2AX cells was determined. Green and blue colors indicate γH2AX and Hoechst, respectively, and scale bars represent 10 μm. The percentage of γH2AX-positive cells was compared using Welch’s *t*-test. **(F)** The combination effect of prexasertib and cisplatin. Cells were treated with each drug concentration for 72 h, and the percentage of growth inhibition is shown relative to untreated controls. Drug synergy was analyzed using CompuSyn software. Error bars represent standard errors of the mean, \**P* < 0.05, \*\**P* < 0.01, and \*\*\**P* < 0.001.

To confirm the prexasertib-induced DNA damage, immunocytochemistry for phospho-H2AX was performed. In SK-UT-1 and SKN cells, a 24-hour exposure to 100 nM prexasertib caused structural abnormalities in the nucleus (Fig. 4E). Moreover, in all cell lines, the percentage of γH2AX-positive cells was significantly increased by prexasertib treatment (in SK-UT-1, SK-LMS-1, and SKN; *P* < 0.001, *P* < 0.001, and *P* < 0.05, respectively, Fig. 4E). In addition, increasing DNA damage is expected to enhance the effect of prexasertib, and therefore, we assessed the combination effect of prexasertib and cisplatin. Cells were treated with a combination of various concentrations of prexasertib and cisplatin, and the synergy was determined using CompuSyn software. As a result, cisplatin synergistically enhanced the effect of prexasertib (Fig. 4F).

### In vivo efficacy of PLK1 and CHEK1 inhibition

Finally, we investigated the *in vivo* efficacy of the inhibitors. SK-UT-1 tumor-bearing mice were treated with either BI 2536 (20 mg/kg or 30 mg/kg) or saline for two weeks after implantation. The mice treated with BI 2536 monotherapy exhibited marked tumor regression (*P* < 0.001, Fig. 5A and Supplementary Fig. S3A). The toxicity was well tolerated, and no mice died due to treatment. When the tumor volume of the control mice reached 2,000 mm^3^, all the mice were sacrificed. The mean tumor weight of the high-dose of BI 2536, low-dose of BI 2536, and control groups were 0.53 g, 0.93 g, and 2.24 g, respectively. Hence, BI 2536 monotherapy significantly decreased the tumor weight in a dose-dependent manner (low-dose and high-dose; *P* < 0.01 and *P* < 0.01, respectively, Fig. 5B).

**Fig. 5.**
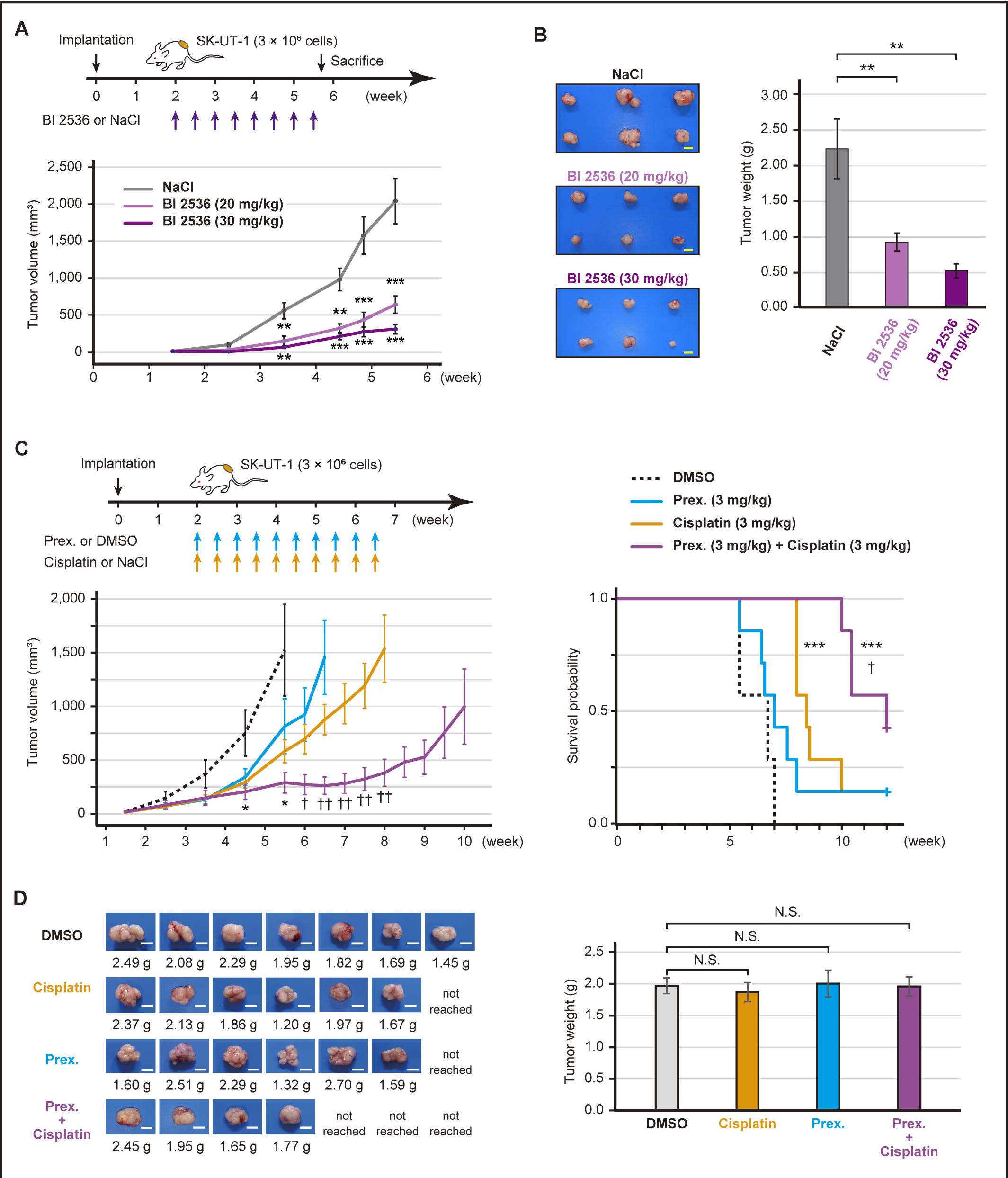
*In vivo* efficacy of BI 2536 or prexasertib. **(A)** Estimated tumor volume of SK-UT-1 tumor-bearing mice treated with either BI 2536 monotherapy or saline (n = 6 per group). High-dose (30 mg/kg), low-dose (20 mg/kg) of BI 2536, or saline was intraperitoneally administered twice a week for four weeks. **(B)** The representative images of tumors and the mean tumor volume of SK-UT-1 tumor-bearing mice treated with either BI 2536 or saline. The mice were sacrificed when the tumors of the control mice reached a volume of 2,000 mm^3^. **(C)** Estimated tumor volume and Kaplan-Meier plot of SK-UT-1 tumor-bearing mice treated with prexasertib and cisplatin in combination (n = 7 per group). Prexasertib (3 mg/kg), cisplatin (3 mg/kg), the combination of prexasertib (3 mg/kg) plus cisplatin (3 mg/kg), or vehicle (DMSO) was intraperitoneally administered twice a week for four weeks. The mice were sacrificed when the tumors reached a volume of 2,000 mm^3^. The tumor volume and weight were compared using Welch’s *t*-test, and survival was compared by a log-rank test. **(D)** The representative images of tumors and the mean tumor volume of SK-UT-1 tumor-bearing mice treated with The prexasertib and cisplatin combination therapy. The tumor weight was compared using Dunnett’s test. The scale bars represent 1 cm. The error bars represent thr standard errors of the mean. \**P* < 0.05, \*\**P* < 0.01, and \*\*\**P* < 0.001 (compared with control mice). †*P* < 0.05 and ††*P* < 0.01 (compared with cisplatin-treated mice), N.S., not significant.

Combination therapy is an effective strategy for increasing therapeutic effects and reducing adverse events. We assessed the anti-cancer effect of the combination of prexasertib (3 mg/kg) with cisplatin (3 mg/kg). The prexasertib monotherapy showed no considerable tumor regression effect, whereas cisplatin monotherapy significantly prolonged survival compared with the DMSO treatment (*P* < 0.001, Fig. 5C). When combined with prexasertib and cisplatin, the anti-cancer effect was enhanced, and marked growth inhibition was observed. Thus, compared with DMSO treatment, the combination therapy significantly reduced the tumor volume (*P* < 0.05) and prolonged survival (*P* < 0.001, Fig. 5C and Supplementary Fig. S3B). In the combination therapy group, the tumors of three mice did not reach a volume of 2,000 mm^3^ within 12 weeks. The tumor weights of sacrificed mice were not significantly different between the groups, indicating that the experiment was conducted fairly (Fig. 5C). Moreover, the survival period in the combination group was significantly longer compared with that in the cisplatin monotherapy group (p < 0.05, Fig. 5C&5D). Therefore, BI 2536 monotherapy and prexasertib plus cisplatin combination therapy caused significant tumor regression in a ULMS mouse model.

## Discussion

In the present study, we identified the key regulators involved in the cell cycle and DNA damage response that activated in ULMS. The regulators, including PLK1 and CHEK1, are potential therapeutic targets, and their selective inhibitors showed outstanding antitumor effects both *in vitro* and *in vivo*.

ULMS is one of the most aggressive gynecological malignancies; therefore, the activation of cell cycle-related genes in ULMS is consistent with this phenotype. Previous reports have also shown the alterations in cell cycle-related signaling pathways in ULMS (9,14). Therefore, the activation of these pathways is a dominant feature of ULMS, and they may represent novel therapeutic targets. For example, Aurora kinase A targeted therapy hindered the growth of ULMS in preclinical models, which prompted a clinical trial of alisertib, an Aurora kinase inhibitor (14,15). However, in the phase II trial, which included 23 recurrent/persistent ULMS patients, the median PFS was 1.7 months; thus, alisertib did not demonstrate clinically significant single-agent activity (15). Therefore, it is essential to investigate other cell cycle-related target molecules and drugs.

PLK1 is a highly conserved serine/threonine protein kinases and is involved in the regulation of cell division (17,18). PLK1 is overexpressed in various kinds of cancers, and cancer cells often have elevated activity of PLK1 (17,18). Hence, PLK1 has been considered a promising therapeutic target, and BI 2536, a prototype PLK1 inhibitor, was developed (19). BI 2536 induced cell cycle arrest and apoptosis and had a greater anti-cancer effect in the mice models (19-22). Therefore, several clinical trials have been conducted. In the phase II trial for advanced non-small cell lung cancer (NSCLC), BI 2536 monotherapy had modest efficacy, and 4.2% of patients had a partial response (23). However, in the other phase II trials, BI 2536 monotherapy showed limited efficacy in various solid tumors (24-26). Then, volasertib was developed, and phase II trials demonstrated insufficient single-agent activity in metastatic urothelial cancer and NSCLC (27,28). Thus, to maximize the therapeutic effect of PLK inhibitors, several studies have investigated combination therapy. The PLK1 and mTOR targeting therapy induced a synergistic effect in squamous cell carcinoma, and histone deacetylase inhibitors also increased the effect of PLK inhibitors synergistically in prostate cancer cells (29,30). Moreover, an alternative approach would be the exploration of predictive biomarkers for PLK1 inhibition. In NSCLC, more mesenchymal-like cancer cells were more sensitive to PLK1 inhibitors, and PIM1-overexpressing prostate cancer cells were highly sensitive to BI 2536 (31,32). PIM kinases were reported to have a certain role in sarcoma development in an experimental model (33). Therefore, it is expected that ULMS is highly sensitive to PLK1 inhibition, and a suitable combination of drugs is a rational therapeutic strategy.

CHEK1 and CHEK2 are the central regulators of DNA damage response signaling. Briefly, ATR and ATM act as sensors of single-strand and double-strand breaks, respectively, which activate CHEK1 and CHEK2 by phosphorylation. CHEK1 and CHEK2 prevent the removal of phosphates on CDK1 and CDK2 by suppressing CDC25A and CDC25C phosphatases (34). Therefore, activation of CHEK1 and CHEK2 provides the cell time to repair DNA damage. We demonstrated that CHEK1 was more responsible for sarcoma cell proliferation compared with CHEK2. Moreover, previous reports also showed that CHEK1 inhibition causes the inappropriate activation of the CDC25A-CDK2 axis as well as various abnormalities, such as increased double-stranded DNA breaks, the accumulation of aberrant replication fork structures, and the permission to enter the G2/M phase with damaged DNA (35-37). Hence, CHEK1 inhibition may be a novel therapeutic candidate for ULMS. Prexasertib (LY2606368) is an ATP-competitive protein kinase inhibitor with a Ki of 0.9 nmol/L against purified CHEK1. Several studies have shown its excellent antitumor effects in a variety of cancer cells (37-43). Moreover, consistent with our results, a synergistic effect for the combination of prexasertib and cytotoxic drugs or PARP inhibitors has been reported (39-43). In a clinical trial, prexasertib monotherapy demonstrated single-agent activity in heavily pretreated squamous cell carcinoma (44). Moreover, another phase II study also showed the efficacy of prexasertib in *BRCA* wild-type, recurrent high-grade serous ovarian carcinoma (HGSOC), with eight of 24 patients exhibiting partial responses (45). However, the clinical efficacy of prexasertib was modest in advanced *BRCA* wild-type triple-negative breast cancer, despite similar molecular features with HGSOC (46). The posthoc analysis of HGSOC indicated that prexasertib activity might be associated with CCNE1 amplification and overexpression (45). This result is interesting because CCNE1 is amplified and overexpressed in ULMS (9). Therefore, the clinical benefit of prexasertib is highly anticipated in ULMS patients.

According to clinical trials, the toxicity of BI 2536, volasertib, and prexasertib were well tolerable. In patients who administered BI 2536 or volasertib, neutropenia was the most frequently observed adverse event, and about 30%–40% of the patients experienced grade 3 or 4 neutropenia (23-25,27,28). Similarly, hematological adverse events were frequently observed in the patients that were treated with prexasertib, and almost all the patients experienced grade 3 or 4 neutropenia (44-47). Therefore, the hematological toxicity of these drugs should be kept in mind; however, it is important to also note that all clinical trials have concluded the safety profile.

There were several limitations to this study. Firstly, we did not assess the regulatory mechanisms responsible for the activation of the cell cycle in ULMS. This would be interesting as it may lead to the identification of new therapeutic targets. Secondly, we primarily investigated the effect of BI 2536 and prexasertib. However, other cell cycle genes are promising targets for cancer therapy (35). In particular, CDKs inhibition is an attractive treatment strategy based on our results. Therefore, we believe that additional research projects will continue to improve the prognosis of patients with ULMS.

In conclusion, the overexpression of PLK1 and CHEK1 was one hallmark of ULMS. Both BI 2536 and prexasertib strongly induced cell cycle arrest and inhibited the proliferation of ULMS cells. Therefore, PLK1 or CHEK1 inhibition are promising therapeutic strategies that might improve clinical outcomes for ULMS.

## Data Availability

https://www.ncbi.nlm.nih.gov/geo/query/acc.cgi?acc=GSE185543

## Acknowledgments

Research reported in this publication was supported by the Program for Promoting the Enhancement of Research Universities as young researcher units for the advancement of new and undeveloped fields at Nagoya University. We thank the National Cancer Center Biobank for providing biological resources. We received technical support from Yuko Fujiwara at the Laboratory of Molecular Carcinogenesis, National Cancer Center Research Institute. Moreover, we thank Enago (www.enago.jp) for the English language review.

## Financial support

This study was supported by JSPS KAKENHI Grant Numbers 21H02721, 21H03075, and 21K16789. Moreover, YOKOYAMA Foundation for Clinical Pharmacology (YRY-2115), Japan Research Foundation for Clinical Pharmacology (2021A18), and Foundation for Promotion of Cancer Research in Japan supported as well.

## Figure legends

**Supplementary Figure S1. The analysis of three GEO datasets (A)** The Venn diagram showing common gene symbols in GSE36610, GSE64763, and GSE68295. **(B)** The volcano plot showing significant DEGs between LMSs and myometrium in each dataset. The adjusted *P* values for each gene were calculated by the Wald test in DESeq2.

**Supplementary Figure S2. The inhibitory effects of selective inhibitors (A)** The effect of an approved drug, pazopanib. **(B)** The effect of dinaciclib, flavopiridol, AT9283, tozasertib, JNJ-7706621, and BAY 1217389. Dinaciclib and flavopiridol are pan-CDK inhibitors. AT9283 is a JAK2/3 and Aurora A/B inhibitor, and tozasertib is a pan-Aurora inhibitor. JNJ-7706621 is a pan-CDK and potent Aurora A/B inhibitor, and BAY 1217389 is a TTK inhibitor. The cells were treated with each inhibitor for 72 h. Red, green, and blue represent SK-UT-1, SK-LMS-1, and SKN, respectively. Experiments were performed in triplicate and repeated three times, and IC50 and drug dose-response curves were calculated in GraphPad Prism 7.

**Supplementary Figure S3. Tumor growth in SK-UT-1 tumor-bearing mice (A)** The estimated tumor volume of SK-UT-1 tumor-bearing mice treated with either BI 2536 monotherapy or saline. High-dose (30 mg/kg), low-dose (20 mg/kg) of BI 2536, or saline were intraperitoneally administered twice a week for four weeks. **(B)** The estimated tumor volume of SK-UT-1 tumor-bearing mice treated with prexasertib and cisplatin combination therapy. Prexasertib (3 mg/kg), cisplatin (3 mg/kg), the combination of prexasertib (3 mg/kg) plus cisplatin (3 mg/kg), or vehicle (DMSO) was intraperitoneally administered twice a week for four weeks.

## Notes

**Additional information** Financial support: This study was supported by JSPS KAKENHI Grant Numbers 21H02721, 21H03075, and 21K16789. Moreover, YOKOYAMA Foundation for Clinical Pharmacology (YRY-2115), Japan Research Foundation for Clinical Pharmacology (2021A18), and Foundation for Promotion of Cancer Research in Japan supported as well.

A conflict of interest disclosure statement: The authors declare no potential conflicts of interest.

### Competing Interest Statement

The authors have declared no competing interest.

### Author Declarations

The ethics committee of the National Cancer Center (Tokyo, Japan) gave ethical approval for this work (approval No. 2020-160).

## References

1. Roberts ME, Aynardi JT, Chu CS. Uterine leiomyosarcoma: A review of the literature and update on management options. Gynecol Oncol 2018;151(3):562–72.

2. George S, Serrano C, Hensley ML, Ray-Coquard I. Soft tissue and uterine leiomyosarcoma. J Clin Oncol 2018;36(2):144–50.

3. Skorstad M, Kent A, Lieng M. Uterine leiomyosarcoma - incidence, treatment, and the impact of morcellation. A nationwide cohort study. Acta Obstet Gynecol Scand 2016;95(9):984–90.

4. Seagle BL, Sobecki-Rausch J, Strohl AE, Shilpi A, Grace A, Shahabi S. Prognosis and treatment of uterine leiomyosarcoma: A National Cancer Database study. Gynecol Oncol 2017;145(1):61–70.

5. Abeler VM, Røyne O, Thoresen S, Danielsen HE, Nesland JM, Kristensen GB. Uterine sarcomas in Norway. A histopathological and prognostic survey of a total population from 1970 to 2000 including 419 patients. Histopathology 2009;54(3):355–64.

6. Hensley ML, Patel SR, von Mehren M, Ganjoo K, Jones RL, Staddon A, et al. Efficacy and safety of trabectedin or dacarbazine in patients with advanced uterine leiomyosarcoma after failure of anthracycline-based chemotherapy: Subgroup analysis of a phase 3, randomized clinical trial. Gynecol Oncol 2017;146(3):531–7.

7. Blay JY, Schöffski P, Bauer S, Krarup-Hansen A, Benson C, D’Adamo DR, et al. Eribulin versus dacarbazine in patients with leiomyosarcoma: subgroup analysis from a phase 3, open-label, randomised study. Br J Cancer 2019;120(11):1026–32.

8. Benson C, Ray-Coquard I, Sleijfer S, Litière S, Blay JY, Le Cesne A, et al. Outcome of uterine sarcoma patients treated with pazopanib: A retrospective analysis based on two European Organisation for Research and Treatment of Cancer (EORTC) Soft Tissue and Bone Sarcoma Group (STBSG) clinical trials 62043 and 62072. Gynecol Oncol 2016;142(1):89–94.

9. Cuppens T, Moisse M, Depreeuw J, Annibali D, Colas E, Gil-Moreno A, et al. Integrated genome analysis of uterine leiomyosarcoma to identify novel driver genes and targetable pathways. Int J Cancer 2018;142(6):1230–43.

10. Hensley ML, Chavan SS, Solit DB, Murali R, Soslow R, Chiang S, et al. Genomic landscape of uterine sarcomas defined through prospective clinical sequencing. Clin Cancer Res 2020;26(14):3881–8.

11. Astolfi A, Nannini M, Indio V, Schipani A, Rizzo A, Perrone AM, et al. Genomic database analysis of uterine leiomyosarcoma mutational profile. Cancers (Basel) 2020;12(8).

12. Choi J, Manzano A, Dong W, Bellone S, Bonazzoli E, Zammataro L, et al. Integrated mutational landscape analysis of uterine leiomyosarcomas. Proc Natl Acad Sci U S A 2021;118(15).

13. Mas A, Alonso R, Garrido-Gómez T, Escorcia P, Montero B, Jiménez-Almazán J, et al. The differential diagnoses of uterine leiomyomas and leiomyosarcomas using DNA and RNA sequencing. Am J Obstet Gynecol 2019;221(4):320.e1–.e23.

14. Shan W, Akinfenwa PY, Savannah KB, Kolomeyevskaya N, Laucirica R, Thomas DG, et al. A small-molecule inhibitor targeting the mitotic spindle checkpoint impairs the growth of uterine leiomyosarcoma. Clin Cancer Res 2012;18(12):3352–65.

15. Hyman DM, Sill MW, Lankes HA, Piekarz R, Shahin MS, Ridgway MR, et al. A phase 2 study of alisertib (MLN8237) in recurrent or persistent uterine leiomyosarcoma: An NRG Oncology/Gynecologic Oncology Group study 0231D. Gynecol Oncol 2017;144(1):96–100.

16. Bray NL, Pimentel H, Melsted P, Pachter L. Near-optimal probabilistic RNA-seq quantification. Nat Biotechnol 2016;34(5):525–7.

17. Schöffski P. Polo-like kinase (PLK) inhibitors in preclinical and early clinical development in oncology. Oncologist 2009;14(6):559–70.

18. Archambault V, Lépine G, Kachaner D. Understanding the Polo Kinase machine. Oncogene 2015;34(37):4799–807.

19. Steegmaier M, Hoffmann M, Baum A, Lénárt P, Petronczki M, Krssák M, et al. BI 2536, a potent and selective inhibitor of polo-like kinase 1, inhibits tumor growth in vivo. Curr Biol 2007;17(4):316–22.

20. Ding Y, Huang D, Zhang Z, Smith J, Petillo D, Looyenga BD, et al. Combined gene expression profiling and RNAi screening in clear cell renal cell carcinoma identify PLK1 and other therapeutic kinase targets. Cancer Res 2011;71(15):5225–34.

21. Grinshtein N, Datti A, Fujitani M, Uehling D, Prakesch M, Isaac M, et al. Small molecule kinase inhibitor screen identifies polo-like kinase 1 as a target for neuroblastoma tumor-initiating cells. Cancer Res 2011;71(4):1385–95.

22. Maire V, Némati F, Richardson M, Vincent-Salomon A, Tesson B, Rigaill G, et al. Polo-like kinase 1: a potential therapeutic option in combination with conventional chemotherapy for the management of patients with triple-negative breast cancer. Cancer Res 2013;73(2):813–23.

23. Sebastian M, Reck M, Waller CF, Kortsik C, Frickhofen N, Schuler M, et al. The efficacy and safety of BI 2536, a novel Plk-1 inhibitor, in patients with stage IIIB/IV non-small cell lung cancer who had relapsed after, or failed, chemotherapy: results from an open-label, randomized phase II clinical trial. J Thorac Oncol 2010;5(7):1060–7.

24. Mross K, Dittrich C, Aulitzky WE, Strumberg D, Schutte J, Schmid RM, et al. A randomised phase II trial of the Polo-like kinase inhibitor BI 2536 in chemo-naïve patients with unresectable exocrine adenocarcinoma of the pancreas - a study within the Central European Society Anticancer Drug Research (CESAR) collaborative network. Br J Cancer 2012;107(2):280–6.

25. Awad MM, Chu QS, Gandhi L, Stephenson JJ, Govindan R, Bradford DS, et al. An open-label, phase II study of the polo-like kinase-1 (Plk-1) inhibitor, BI 2536, in patients with relapsed small cell lung cancer (SCLC). Lung Cancer 2017;104:126–30.

26. Schöffski P, Blay JY, De Greve J, Brain E, Machiels JP, Soria JC, et al. Multicentric parallel phase II trial of the polo-like kinase 1 inhibitor BI 2536 in patients with advanced head and neck cancer, breast cancer, ovarian cancer, soft tissue sarcoma and melanoma. The first protocol of the European Organization for Research and Treatment of Cancer (EORTC) Network Of Core Institutes (NOCI). Eur J Cancer 2010;46(12):2206–15.

27. Stadler WM, Vaughn DJ, Sonpavde G, Vogelzang NJ, Tagawa ST, Petrylak DP, et al. An open-label, single-arm, phase 2 trial of the Polo-like kinase inhibitor volasertib (BI 6727) in patients with locally advanced or metastatic urothelial cancer. Cancer 2014;120(7):976–82.

28. Ellis PM, Leighl NB, Hirsh V, Reaume MN, Blais N, Wierzbicki R, et al. A Randomized, Open-Label Phase II Trial of Volasertib as Monotherapy and in Combination With Standard-Dose Pemetrexed Compared With Pemetrexed Monotherapy in Second-Line Treatment for Non-Small-Cell Lung Cancer. Clin Lung Cancer 2015;16(6):457–65.

29. Wissing MD, Mendonca J, Kortenhorst MS, Kaelber NS, Gonzalez M, Kim E, et al. Targeting prostate cancer cell lines with polo-like kinase 1 inhibitors as a single agent and in combination with histone deacetylase inhibitors. Faseb j 2013;27(10):4279–93.

30. Liu TT, Yang KX, Yu J, Cao YY, Ren JS, Hao JJ, et al. Co-targeting PLK1 and mTOR induces synergistic inhibitory effects against esophageal squamous cell carcinoma. J Mol Med (Berl) 2018;96(8):807–17.

31. Ferrarotto R, Goonatilake R, Yoo SY, Tong P, Giri U, Peng S, et al. Epithelial-Mesenchymal Transition Predicts Polo-Like Kinase 1 Inhibitor-Mediated Apoptosis in Non-Small Cell Lung Cancer. Clin Cancer Res 2016;22(7):1674–86.

32. van der Meer R, Song HY, Park SH, Abdulkadir SA, Roh M. RNAi screen identifies a synthetic lethal interaction between PIM1 overexpression and PLK1 inhibition. Clin Cancer Res 2014;20(12):3211–21.

33. Narlik-Grassow M, Blanco-Aparicio C, Cecilia Y, Peregrina S, Garcia-Serelde B, Muñoz-Galvan S, et al. The essential role of PIM kinases in sarcoma growth and bone invasion. Carcinogenesis 2012;33(8):1479–86.

34. Lin AB, McNeely SC, Beckmann RP. Achieving precision death with cell-cycle inhibitors that target DNA replication and repair. Clin Cancer Res 2017;23(13):3232–40.

35. Otto T, Sicinski P. Cell cycle proteins as promising targets in cancer therapy. Nat Rev Cancer 2017;17(2):93–115.

36. Syljuåsen RG, Sørensen CS, Hansen LT, Fugger K, Lundin C, Johansson F, et al. Inhibition of human Chk1 causes increased initiation of DNA replication, phosphorylation of ATR targets, and DNA breakage. Mol Cell Biol 2005;25(9):3553–62.

37. King C, Diaz HB, McNeely S, Barnard D, Dempsey J, Blosser W, et al. LY2606368 causes replication catastrophe and antitumor effects through CHK1-dependent mechanisms. Mol Cancer Ther 2015;14(9):2004–13.

38. Nair J, Huang TT, Murai J, Haynes B, Steeg PS, Pommier Y, et al. Resistance to the CHK1 inhibitor prexasertib involves functionally distinct CHK1 activities in BRCA wild-type ovarian cancer. Oncogene 2020;39(33):5520–35.

39. Parmar K, Kochupurakkal BS, Lazaro JB, Wang ZC, Palakurthi S, Kirschmeier PT, et al. The CHK1 inhibitor prexasertib exhibits monotherapy activity in high-grade serous ovarian cancer models and sensitizes to PARP inhibition. Clin Cancer Res 2019;25(20):6127–40.

40. Mani C, Jonnalagadda S, Lingareddy J, Awasthi S, Gmeiner WH, Palle K. Prexasertib treatment induces homologous recombination deficiency and synergizes with olaparib in triple-negative breast cancer cells. Breast Cancer Res 2019;21(1):104.

41. Lowery CD, Dowless M, Renschler M, Blosser W, VanWye AB, Stephens JR, et al. Broad spectrum activity of the checkpoint kinase 1 inhibitor prexasertib as a single agent or chemopotentiator across a range of preclinical pediatric tumor models. Clin Cancer Res 2019;25(7):2278–89.

42. Sen T, Tong P, Stewart CA, Cristea S, Valliani A, Shames DS, et al. CHK1 inhibition in small-cell lung cancer produces single-agent activity in biomarker-defined disease subsets and combination activity with cisplatin or olaparib. Cancer Res 2017;77(14):3870–84.

43. Heidler CL, Roth EK, Thiemann M, Blattmann C, Perez RL, Huber PE, et al. Prexasertib (LY2606368) reduces clonogenic survival by inducing apoptosis in primary patient-derived osteosarcoma cells and synergizes with cisplatin and talazoparib. Int J Cancer 2020;147(4):1059–70.

44. Hong DS, Moore K, Patel M, Grant SC, Burris HA, 3rd, William WN, Jr., et al. Evaluation of prexasertib, a checkpoint kinase 1 inhibitor, in a phase Ib study of patients with squamous cell carcinoma. Clin Cancer Res 2018;24(14):3263–72.

45. Lee JM, Nair J, Zimmer A, Lipkowitz S, Annunziata CM, Merino MJ, et al. Prexasertib, a cell cycle checkpoint kinase 1 and 2 inhibitor, in BRCA wild-type recurrent high-grade serous ovarian cancer: a first-in-class proof-of-concept phase 2 study. Lancet Oncol 2018;19(2):207–15.

46. Gatti-Mays ME, Karzai FH, Soltani SN, Zimmer A, Green JE, Lee MJ, et al. A phase II single arm pilot study of the CHK1 inhibitor prexasertib (LY2606368) in BRCA wild-type, advanced triple-negative breast cancer. Oncologist 2020.

47. Iwasa S, Yamamoto N, Shitara K, Tamura K, Matsubara N, Tajimi M, et al. Dose-finding study of the checkpoint kinase 1 inhibitor, prexasertib, in Japanese patients with advanced solid tumors. Cancer Sci 2018;109(10):3216–23.

