## Supplementary figures and images for "Integrative transcriptomics analysis for uterine leiomyosarcoma identifies aberrant activation of cell cycle-dependent kinases and their potential therapeutic significance"

### Supplementary Figure S1

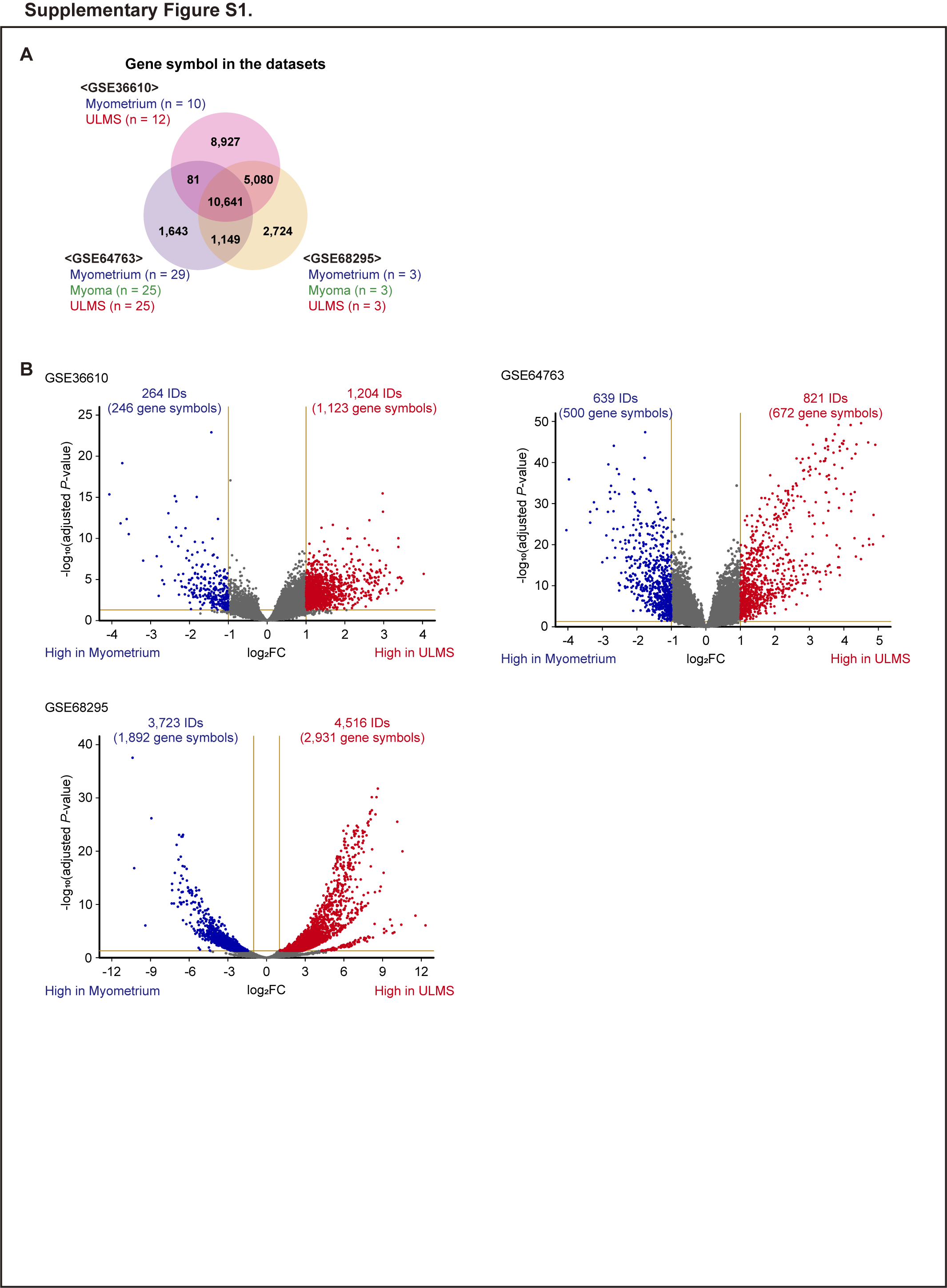

### Supplementary Figure S2

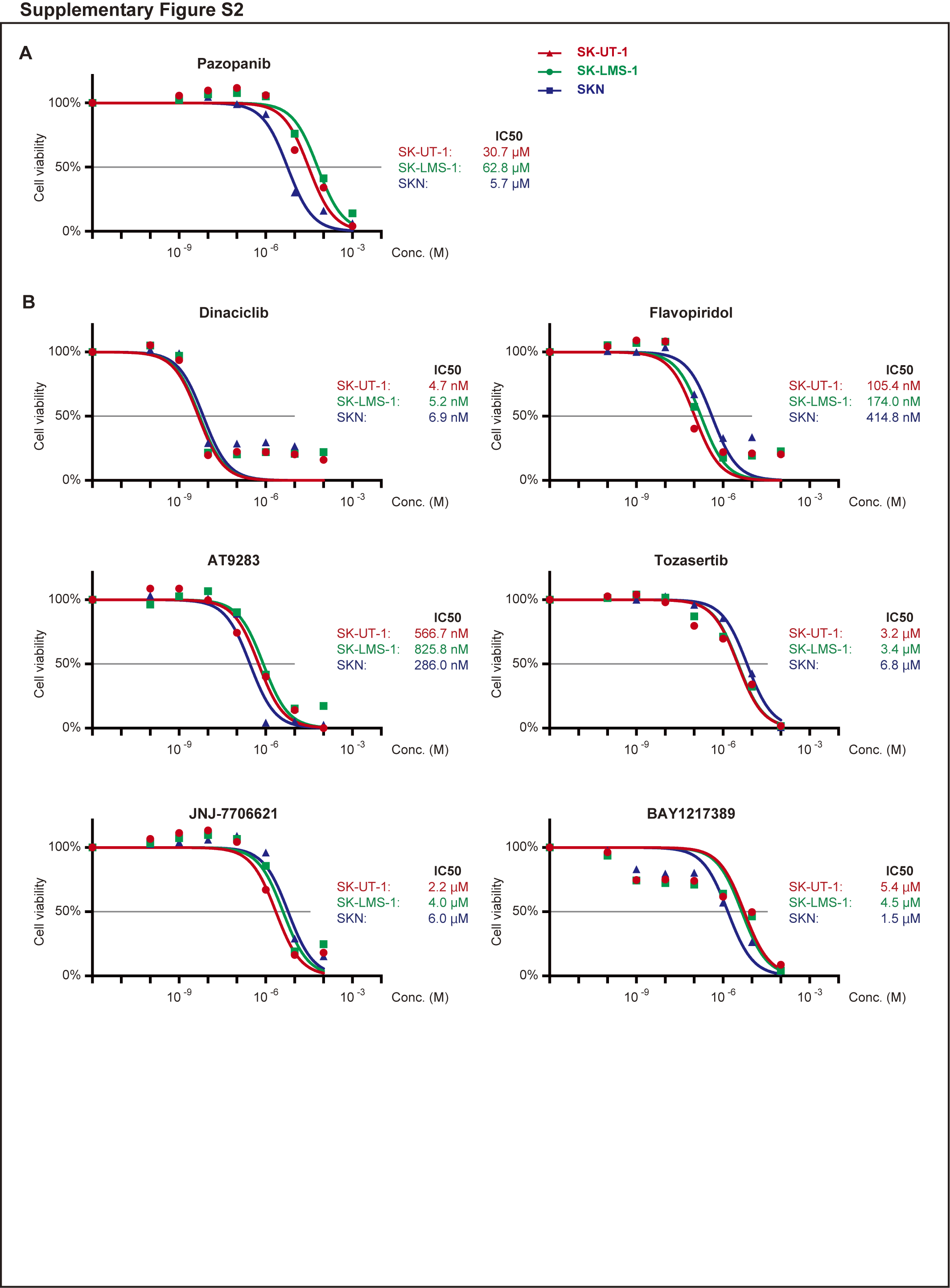

### Supplementary Figure S3

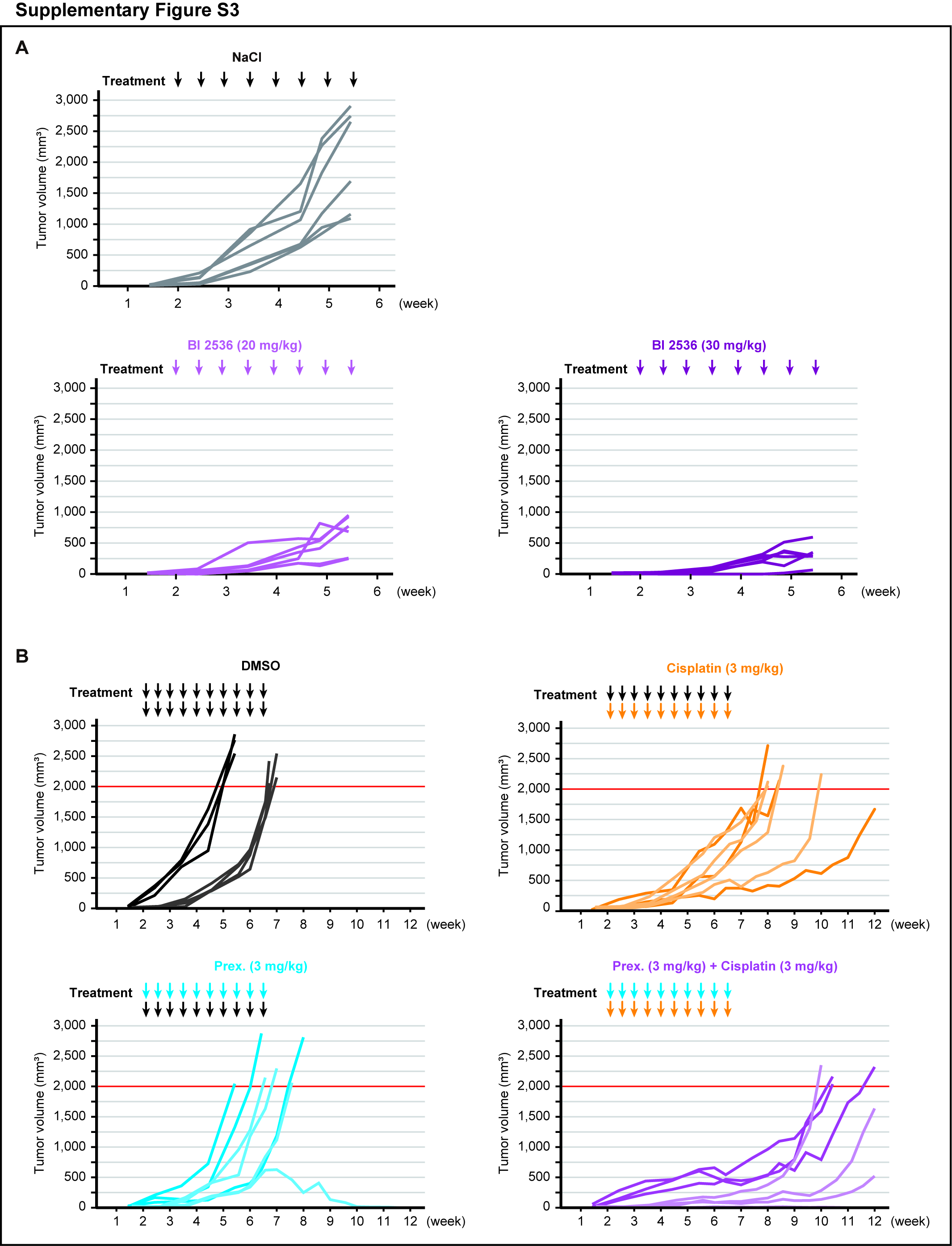
